# Keep the hospital clean: diagnostic performance of ten different molecular and culture-based methods to detect *Candidozyma auris*

**DOI:** 10.1101/2025.01.18.25320756

**Authors:** Koos Korsten, Bert Gerrits van den Ende, Rick D. Pique, Ferry Hagen, Karin van Dijk

**Affiliations:** Department of Medical Microbiology and Infection Prevention, Amsterdam University Medical Center, location AMC, Amsterdam, the Netherlands; Westerdijk Fungal Biodiversity Institute, Utrecht, The Netherlands; Institute for Biodiversity and Ecosystem Dynamics, University of Amsterdam, Amsterdam, The Netherlands; Department of Medical Microbiology, University Medical Center Utrecht, Utrecht, The Netherlands

## Abstract

**Rationale:** *Candidozyma auris* (formerly *Candida auris*) is a globally emerging potentially multi-drug resistant human pathogenic yeast. To detect *C. auris* we aimed to compare different culture-, and molecular-based methods.

**Methods:** Rectal swabs routinely collected in clinical care were spiked with different concentrations of *C. auris*. Co-infection/colonization was mimicked by spiking part of these samples with other pathogenic *Candida* species. Spiked materials were cultured at 37°C or 42°C using CHROMagar Candida and CHROMagar CandidaPlus plates. In parallel, samples were incubated in a dulcitol salt enrichment broth. Additionally, we compared seven in-house and commercial molecular tests on the direct material and from the broth one day after inoculation.

**Results:** Culture-based methods showed sensitivities up to 100% within 48 hours of incubation, although sensitivity decreased as low as 44% at lower concentrations (≤50 CFU per inoculum), in the presence of an abundance of other species and at higher temperature (42°C). Incubation at 42°C made visual identification possible since other species with similar colony morphologies did not grow at this temperature. No added value of using the dulcitol salt enrichment broth was found. qPCR on direct materials was highly sensitive and specific (both up to 100%) but major differences between various molecular tests were observed.

**Conclusion:** We showed that both culture-based and molecular methods are sensitive for diagnosing *C. auris*. The clinical setting (routine screening versus an outbreak), local prevalence and the load in those that carry or are infected by *C. auris* are important factors to consider when determining which diagnostic tests should be employed.

## Introduction

The yeast *Candidozyma auris* (formerly *Candida auris*, taxonomically reassigned to the genus *Candidozyma* together with other members of the *Candida haemuli* clade [1]), is emerging worldwide and has received a critical priority listing by the World Health Organization [2]. *C. auris* is primarily responsible for healthcare associated infections in those who are critically ill or those with underlying medical conditions such as immunocompromised patients. This pathogen provides a challenge for the healthcare settings because of potential of increased antifungal resistance and the prolonged colonization in both patients and on fomites which can ease transmission and result in persistence despite the use of standard healthcare disinfectants [2, 3]. It is therefore important to implement surveillance to accomplish adequate infection prevention measures and limit the spread of this pathogen. Different diagnostic methods have been evaluated for the isolation and identification of *C. auris*, each with its own strengths and limitations. *C. auris* will grow on standard media but might be overgrown by other microorganisms for which chromogenic culture media can be used to aid in visual identification [4–6]. Another method to recover *C. auris* is by using a selective enrichment broth such as those with high salinity and dulcitol as a carbon source [3, 6]. Molecular methods can also be used for quick identification of *C. auris.* These include various commercial and in-house developed qPCRs as well as sequencing of ribosomal DNA genes (ITS and D1/D2). These methods are accurate, relatively quick as compared to culture methods, but are expensive and could represent “dead” yeasts [7, 8]. Matrix-assisted laser desorption/ionization – time of flight mass spectrometry (MALDI-TOF MS) identifies *C. auris* accurately and is cheap but is dependent on culture growth [8]. The aim of the current study is to compare the different diagnostic methods for the identification of *C. auris* in order to identify a valid screening method to establish a *C. auris* diagnostic workflow.

## Methods

In this study we compared ten diagnostic testing methods using clinical materials spiked with *C. auris* from the five currently known circulating clades. We compared seven molecular tests and three culture-based methods including the CHROMagar Candida (CC, CHROMagar, Paris, France) and CHROMagar Candida Plus (CCP, CHROMagar) incubated at 37°C and 42°C. In parallel, we inoculated a 2.5 ml dulcitol 10% NaCl enrichment broth [3]. This broth combines a high salinity and alternative carbon source (dulcitol) to selectively grow *C. auris*.

### Strains

In order to spike the clinical materials we used nine *C. auris* strains, two strains per clade, representing the four clades that are specific to each geographic region and one strain representing the fifth clade [4]. Strains came from reputable international culture collections, including the ‘Centraalbureau voor Schimmelcultures’ (CBS) yeast collection hosted at the Westerdijk Fungal Biodiversity Institute (Utrecht, The Netherlands). We additionally used two mixtures of other *Candida* species to mimic coinfection/colonization and to test the sensitivity and specificity of the diagnostic methods. The ‘genetic’ mixture included the following *Candida* species, closely related to *C. auris*: *Candidozyma haemuli, Candidozyma pseudohaemuli*, *Candidozyma duobushaemuli*, *Candidozyma khanbhai* and *Candidozyma vulturna (all formerly Candida species)*. The ‘common’ mixture, included frequently identified *Candida* species in clinical materials: *Candida albicans*, *Candida parapsilosis*, *Candida tropicalis, Nakaseomyces glabratus (*formerly *Candida glabrata*) and *Pichia kudriavzevii (*formerly *Candida krusei*). The list of strains used can be found in the Supplemental Table S1.

### Sample preparation

Rectal swabs were collected from ICU patients and patients with hematologic malignancies who were routinely screened for carriage of yeasts and Gram-negative bacteria using eSwabs (Copan Diagnostics, Murrieta, CA, U.S.A.). We selected samples that were negative for both these entities. The eSwab medium was spiked at different concentrations using the Cellometer cell counter (Nexcelom Bioscience, Lawrence, MA, U.S.A.) to standardize concentrations. Before spiking, strains were sub-cultured for fresh growth on yeast peptone glucose agar at 35°C.

To test the sensitivity, we spiked swabs only with *C. auris* in concentrations of 10 CFU (n=9) and 100 CFU (n=9) per standard inoculum (50µl). Secondly, we spiked swabs with *C. auris* in concentrations of 50 CFU (n=9) and 150 CFU (n=9) per inoculum and subsequently added the mixtures of other *Candida* species (in concentrations of 50 CFU per species per inoculum). For validation of the specificity of the qPCRs we included two samples that were only spiked with either of the mixtures of other *Candida* strains (in concentration of 50 CFU per species per inoculum) and lastly included a negative control (only eSwab medium). This resulted in 18 samples with only *C. auris*, 18 samples with *C. auris* and other *Candida* species, one sample with the ‘genetic’ mixture, one sample with the ‘common’ mixture and one negative control.

### Study procedures

The study procedures are visualized in Figure 1. After the rectal swabs were spiked and vortexed, 500μL of the directly spiked sample was collected for molecular testing. Each sample was streaked on both the regular CHROMagar Candida (CC), and the CHROMagar Candida Plus (CCP) plates using an inoculum of 50µl. Plates were prepared from the dry powder as per the manufacturer’s instructions and equilibrated to room temperature in the dark before use. Plates were incubated at either 37°C or 42°C for 48h before recording their physical appearance. Visual identification of *C. auris* suspected growth was determined and confirmed using MALDI-TOF MS (Bruker, Bremen, Germany). The 10% NaCl dulcitol broth was in-house prepared according to previous literature [3] inoculated using 50 µl of the spiked sample and was incubated for four days at 40°C. After day one we collected 500μL of broth for PCR. The broth was inspected on a daily basis for four days for visual growth. Upon visual growth, Sabouraud dextrose agar plates were inoculated using 50µl of the broth medium and an additional 500μL of medium was collected for molecular testing. Nucleic acids purification was performed for all molecular assays from the 500μL of swab medium using the MagNA Pure 96 in combination with the MagNA Pure 96 DNA and Viral Nucleic Acid Small Volume Kit (all from Roche Diagnostics, Almere, The Netherlands).

**Figure 1.**
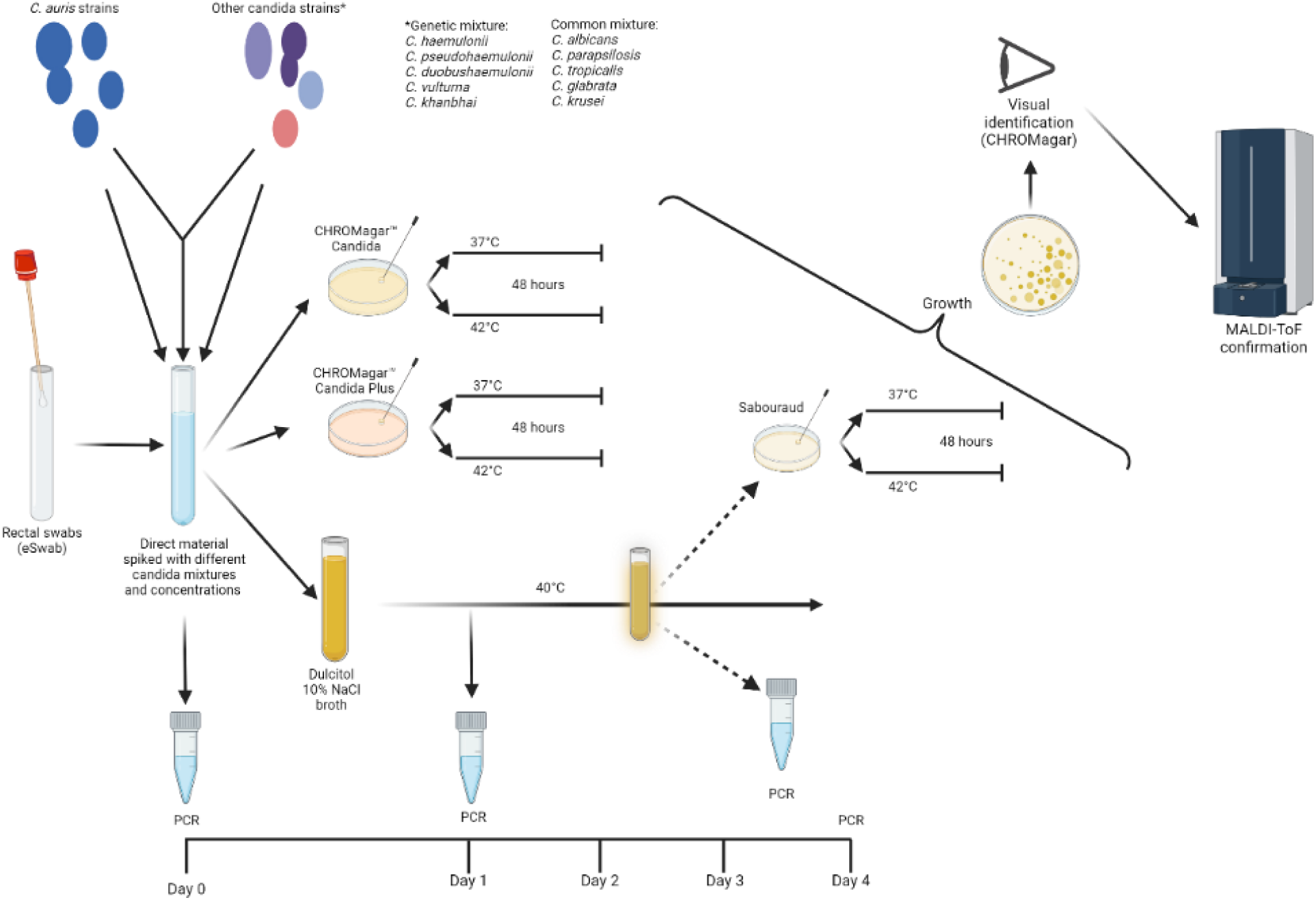
Schematic overview of study procedures.

For molecular testing, we compared seven different qPCRs including three commercially developed; the AurisID (OLM Diagnostics, Braintree, United Kingdom) [9], the *C. auris* AltoStar (Altona Diagnostics, Hamburg, Germany) and the *C. auris* screening assay (Pathonostics, Maastricht, The Netherlands). All commercial PCRs were performed as per the manufacturer’s instructions. We also compared four in-house developed qPCRs. This included the qPCR assays by Leach et al. (2018, [10]), Leonhard et al. (2024, [11]), and the duplex qPCR assay that targets with one primer/probe set the 15 species in the *C. haemuli* species complex [12] and the *C. auris* IDCARD [12] both developed within the Westerdijk Fungal Biodiversity Institute, Utrecht, The Netherlands. Detailed information about the PCRs can be found in supplemental Table S2. All in-house developed tests were performed in volumes of 20µl containing 10µl 2X SensiFast NO-ROX (Meridian Bioscience, Memphis, TE, U.S.A.), final concentrations of 500nM of each primer and 200nM of each probe (all obtained from IDT, San Diego, CA, U.S.A.) in a volume of 2µl, and 8µl sample. All qPCRs were performed on the LC480-II platform of Roche using the settings provided by the manufactures. For the in-house assays the 3-dye filter setting was used (FAM-HEX-Cy5) with the program 10min 95°C, 45 cycli 1sec 95°C, 12sec 60°C (followed by fluorescence measurement), and a 30sec 40°C step to cool down the plate.

### Statistical analysis

We calculated the analytical sensitivity and specificity of the culture and molecular methods in an experimental setting. We determined the accuracy of the molecular results in which accuracy (%) = (true positive + true negative)*100 / total. Estimates and 95% confidence intervals for analytical test performance were calculated using Clopper-Pearson (Exact) method using the epiR package [13]. Data were analyzed in R version 4.0.3.

## Results

### Culture analysis

In samples spiked only with *C. auris*, we observed visible growth after 48h incubation in 29/36 (81%) of CHROMagar Candida (CC) and 30/36 (83%) of the CHROMagar Candida Plus (CCP) plates (Table 1). All plates without visible growth were inoculated with a low concentration of *C. auris* (10 CFU per 50µl). We did not observe an effect of temperature on growth. The plates that were inoculated with a concentration of 100 CFU all grew *C. auris* at both 37°C and 42°C (100% sensitivity). Colony morphology was as expected with cream-pinkish colonies without a halo on the CC agar, and cream-white to cream-pink colonies with a blue-green halo on the CCP agar (Figure 2).

**Figure 2.**
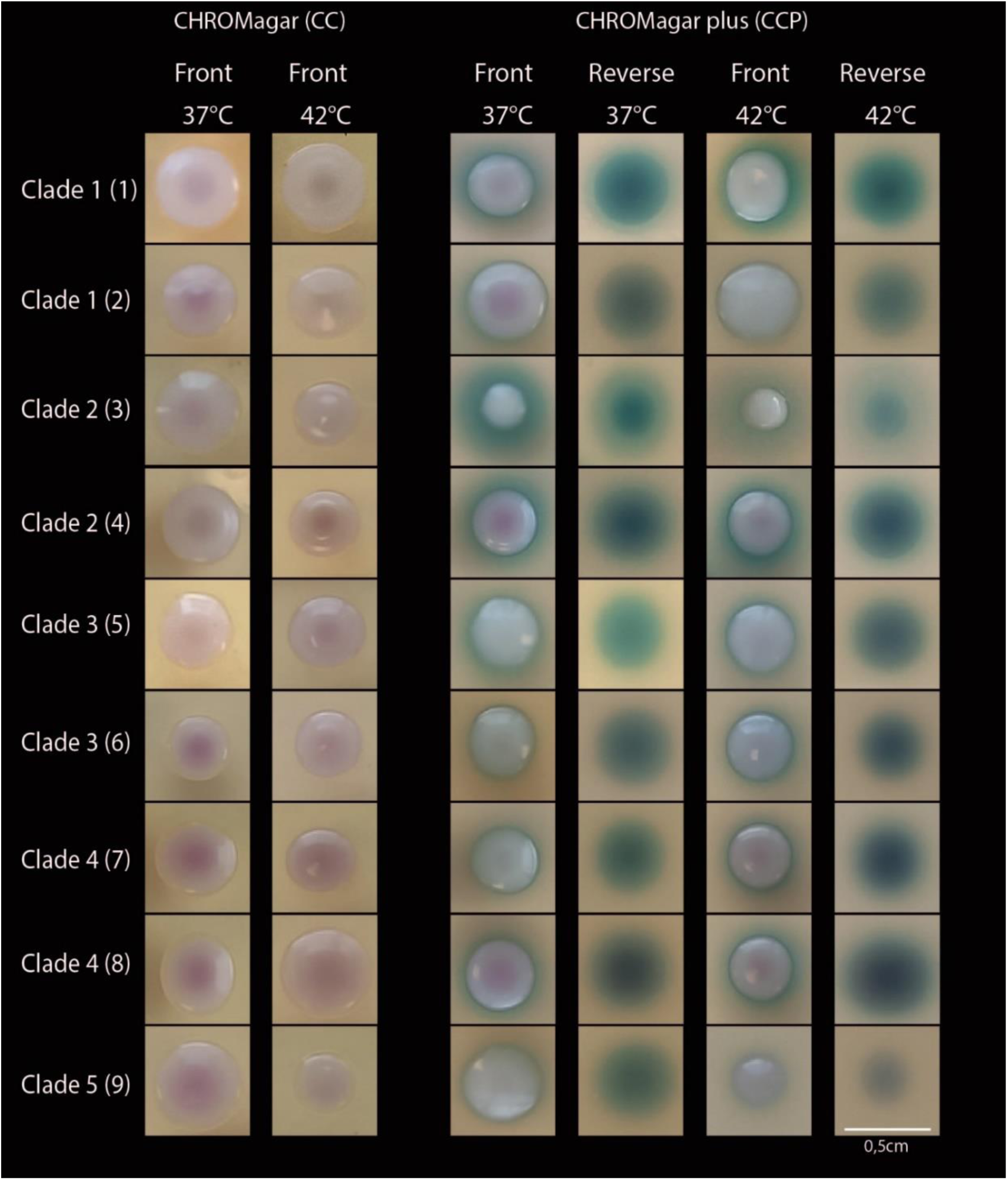
Colony morphology of *C. auris* after 48 hours of incubation. (1) CDC-AR0387 and (2) CDC-AR0388 (clade I, South-Asia); (3) CDC-AR0381 and (4) CDC-AR0382 (clade II, East-Asia; India); (5) CDC-AR0383 and (6) CDC-AR0384 (clade III, Africa); (7) CDC-AR0385 and (8) CDC-AR0386 (clade IV, South-America); (9) CDC-AR1097 (clade V, Iran)

**Table 1.** Sensitivity of the CHROMagar plates in samples spiked only with *C. auris*.

| Inoculum (CFU / 50 µl) | Growth on CHROMagar Candida (CC) |  | Growth on CHROMagar Candida Plus (CCP) |  |
| --- | --- | --- | --- | --- |
|  | 37°C | 42°C | 37°C | 42°C |
| 10 | 67% (6/9) | 56% (5/9) | 67% (6/9) | 67% (6/9) |
| 100 | 100% (9/9) | 100% (9/9) | 100% (9/9) | 100% (9/9) |
| Total | 83% (15/18) | 78% (14/18) | 83% (15/18) | 83% (15/18) |

Next we evaluated samples spiked with *C. auris* and either one of the two mixtures of other *Candida* species. After 48h incubation at 37°C we observed growth of *C. auris-*suspected colonies in 94% (17/18) of CC plates and 100% (18/18) of the CCP plates. Definitive visual identification of *C. auris* was difficult due to similarities with the other ‘genetic mixture’ strains (Figure 3). Despite that colonies of *C. auris* were larger, the distinctive blue-green aura was also produced by *C. khanbhai*.

**Figure 3.**
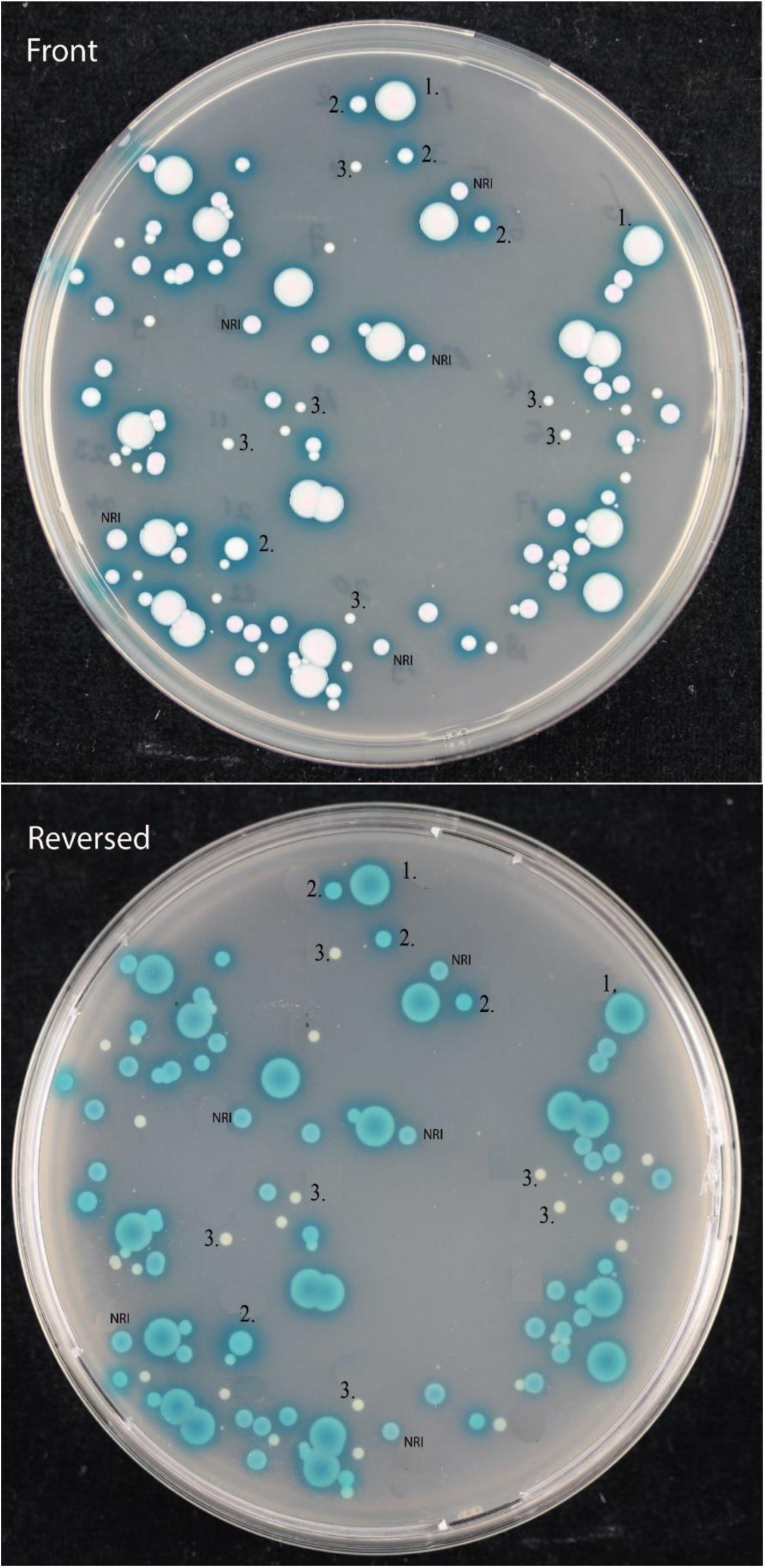
Colony morphology of “genetic mixture” with *C. auris* after 48 hours of incubation. 1. *Candidozyma auris*, 2. *Candidozyma khanbhai* 3. *Candidozyma pseudohaemuli*, NRI no reliable identification (potentially *Candidozyma vulturna*)

At 42°C, we observed only growth from *C. auris, C. albicans* and *P. kudriavzevii* which made visual identification possible based on distinct morphology of the common species (Figure 4). Growth of *C. auris* incubated at 42°C in mixed samples was observed in 66% (12/18) of the CC plates and 72% (13/18) of the CCP plates. When *C. auris* was present in abundance to other species (inoculum of 150 CFU versus 50 CFU for the other species), growth was observed in 89% (8/9) in both the CC and CCP agar. When *C. auris* was present in equal amounts as the other species (all with an inoculum of 50 CFU), only 44% (4/9, CC) and 56% (5/9, CCP) showed visible growth.

**Figure 4.**
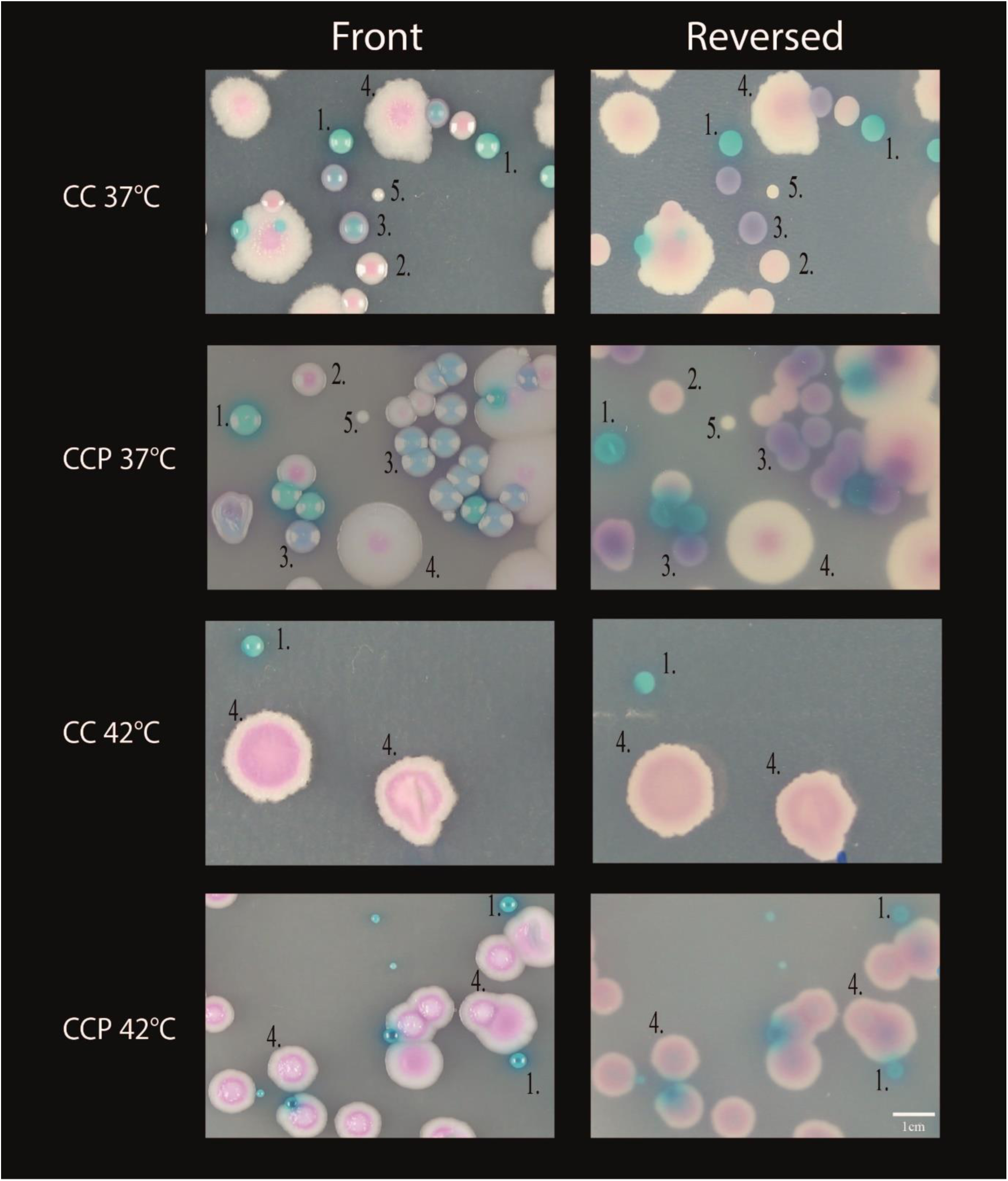
Colony morphology of the “common mixture” after 48 hours incubation. CC = CHROMagar Candida. CCP = CHROMagar Candida Plus. 1. *C. albicans*; 2. *N. glabratus*; 3. *C. tropicalis*; 4. *Pichia kudriavzevii (C. krusei);* 5. *C. parapsilosis*.

None of the inoculated dulcitol broths showed visible growth for four days. Consequently, only the sample for qPCR testing was collected after one day of incubation.

### Molecular analysis

Aliquots for molecular analysis were collected from direct materials after spiking and from the broth after one day of incubation. Results are shown in Table 2. The best performing qPCRs were the in-house PCR by Leonhard [11] and the commercial AltoStar (Altona) which showed a 100% sensitivity and 100% specificity. The AltoStar’s qualitative results were similar although 13 samples had a Ct value of 40 in contrast to only one sample by the qPCR of Leonhard et al. The qPCR by Leach and colleagues showed two false positive results (one in the truly negative control and one in the negative control with only the genetic *Candida* mixture). The qPCR by PathoNostics had the lowest sensitivity with seven false negative results in samples with a *C. auris* load of ≤50 CFU per 50µl. All molecular tests detected higher loads (lower Ct values) for increasing concentrations of *C. auris* (Table 3). Samples collected from the enrichment broth after one day of incubation did not yield additional detection and even showed higher Ct values as compared to samples from the direct materials (Table S3).

**Table 2.** Molecular analysis on direct materials.

| qPCR assay | Sensitivity (95% CI) | Specificity (95% CI) | Accuracy* (95% CI) | TP | FP | TN | FN |
| --- | --- | --- | --- | --- | --- | --- | --- |
| AurisID (OLM) | 83% (67-94) | 100% (29-100) | 85% (69-94) | 30 | 0 | 3 | 6 |
| AltoStar (Altona) | 100% (90-100) | 100% (29-100) | 100% (91-100) | 36 | 0 | 3 | 0 |
| *C. auris* screening assay (PathoNostics) | 81% (64-92) | 100% (29-100) | 82% (66-92) | 29 | 0 | 3 | 7 |
| Leonhard qPCR | 100% (90-100) | 100% (29-100) | 100% (91-100) | 36 | 0 | 3 | 0 |
| Leach qPCR | 94% (81-99) | 33% (1-91%) | 90% (76-97) | 34 | 2 | 1 | 2 |
| *C. auris* IDCARD | 92% (78-98) | 100% (29-100) | 92% (79-98) | 33 | 0 | 3 | 3 |
| *C. haemuli* complex qPCR | 92% (78-98) | 67% (9-99) | 90% (76-97) | 33 | 1** | 2 | 3 |

TP = true positive. FP = false positive. TN = true negative. FN = False negative. \* Accuracy (%) = (true positive + true negative)\*100 / total. \*\* This was not actually a false positive result since this 'negative' sample had no *C. auris* but did include the genetic mixture which should have been positive since this qPCR detects the *C. haemuli* complex.

**Table 3.** Ct values for the samples with different concentrations CFU of *C. auris*.

| PCR | 10 CFU<br>Mean (range) | 50 CFU + mix<br>Mean (range) | 100 CFU<br>Mean (range) | 150 CFU +mix<br>Mean (range) |
| --- | --- | --- | --- | --- |
| AurisID (OLM)* | 38.8 (30.9-33.6) | 33.7 (32.5-35.6) | 30.5 (29.0-32.7) | 29.8 (28.6-31.8) |
| AltoStar (Altona) | 39.5 (37.8-40.0) | 39.6 (38.0-40.0) | 36.6 (34.6-40) | 36.3 (34.9-37.9) |
| *C. auris* screening assay<br>(PathoNostics) | 37.5 (34.5-40.0) | 37.9 (34.9-40.0) | 35.8 (33.0-38.0) | 34.2 (32.8-36.2) |
| Leonhard et al. qPCR | 37.1 (36.5-39.1) | 38.1 (37.0-40.0) | 34.7 (33.6-37.7) | 34.4 (33.4-35.9) |
| Leach et al. qPCR | 36.5 (35.2-38.1) | 37.6 (34.9-40.0) | 33.7 (32.9-35.5) | 33.4 (32.5-35.1) |
| *C. auris* IDCARD | 37.4 (35.6-38.9) | 37.8 (36.3-39.4) | 34.2 (32.8-35.3) | 33.8 (32.5-35.1) |
| *C. haemuli* complex<br>qPCR | 37.7 (36.2-40.0) | 33.2 (31.9-36.3) | 34.2 (33.0-35.1) | 31.5 (31.0-31.9) |

## DISCUSSION

In this study we compared different culture-based, and molecular-based methods to identify *C. auris* in mock-inoculated rectal specimens. Sensitivity of qPCR on direct material ranged from 81-100%. Sensitivity of culture on CHROMagar Candida (CC)/Candida Plus (CCP) ranged from 44-100% and was compromised by low inoculum, the presence of other *Candida* species and/or higher temperature. Visual identification was possible, and most feasible at a temperature of 42°C on both CC and CCP. We did not observe an added value of using the dulcitol salt enrichment broth.

In accordance to previous studies, visual differentiation was possible after two days using the CCP plates due to the distinct halo formation [4, 14]. Early growth however may lack the characteristic pigment/halo production [15]. We observed that *C. khanbhai* produced a similar phenotype including the halo although the colonies were smaller compared to *C. auris*. We could not identify *C. haemuli, C. vulturna* and *C. duobushaemuli* from culture despite that they were included in the sample. Bentz and colleagues [5] showed diminished growth for several *Candida* species in co-culture including *C. haemuli* even at 37°C. *C. vulturna* is not included in the MALDI-TOF MS database in contrast to *C. haemuli* and *C. duobushaemuli. C. vulturna* can also present as a cream-white colony with a blue halo which might have been the colonies without reliable identification from Figure 3. *C. haemuli and C. duobushaemuli* have different morphologies since they do not produce a halo on the CCP agar, but are indistinguishable to *C. auris* on regular CC agar [4]. Since most co-colonization may include common *Candida* species and not include other *Candida* species from the *C. haemuli* complex [16], reliable (visual) identification after 48h is likely possible in a clinical setting on both CC and CCP. Colonies suspected for *C. auris* can subsequently be identified by MALDI-TOF MS. Incubation at higher temperature of 42°C caused selective growth of *C. auris, C. albicans* and *Pichia kudriavzevii.* These species have very distinct morphologies making visual identification feasible. Not all samples of *C. auris* mixed with other *Candida* species showed growth at 42°C (both with the common and genetic mixture). Interactions between the various species might interfere with growth which has been shown for *C. albicans* co-cultured with *N. glabratus* [17] and *C. dubliniensis* [18]. An alternative explanation could have been random sampling variation from the original spiked solution which might have resulted in an inoculum with insufficient viable cells to assure growth. Another factor to consider is the fungal load. We observed a decreased sensitivity in our lowest inoculum group (10 CFU). One might speculate that such a low inoculum is uncommon and perhaps less relevant for clinical transmission. It appears from the study by Zhu and colleagues from a large outbreak in the U.S.A. that patients colonized with *C. auris* harbored large numbers of live cells on skin (axilla/groin, median 10^4^ CFU/swab) and mucosal surfaces (nares, median 10^6^ CFU/swab) [19]. The vast majority of their samples included colonization of at least 100 CFU/swab which might favor incubation at 42°C since it will likely provide sufficient growth and aid the identification process [19]. The knowledge of a positive qPCR prior to culture can facilitate subsequent isolation by laboratory personnel.

In contrast to other studies we did not find an additive value of dulcitol salt enrichment broth [3, 5, 6]. The study by Bentz and colleagues [5] showed 40% reduction in sensitivity in direct culture versus a primary step of using an enrichment broth. However, this benefit might have been caused by an extended time between sample collection and plating of up to 28 days which is not realistic in routine clinical practice. The lack of visible growth in our study could be due to the low volume of the inoculum (50 µl) in a large volume of broth (2.5 ml). Nevertheless, we would have expected visible growth after four days in the groups with a high concentration of *C. auris* which we did not observe.

The strength of this study is that we used clinical materials representative of those collected in clinical care. We spiked these materials using a counting chamber to standardize concentrations of *C. auris* and other genetically-related, and common *Candida* species to mimic various scenarios that could impact diagnostic testing. For validation of the diagnostic method we used all circulating *C. auris* strains from the five known clades. We tested incubation at different temperatures which is an important parameter that could affect specificity of culture-based growth.

This study also has limitations. First, we lacked to include other types of clinical samples (groin, axilla) which are also recommended in screening for *C. auris* [20]. Rectal swab material causes most problems in terms of inhibitory factors that might complicate molecular analysis as compared to other common sample types. Other locations such as the groin or axilla might have a higher incidence of other *Candida* species. By using mixtures of other candida species we mimicked this co-colonization. The high molecular sensitivity and specificity found in our study is therefore reassuring. Secondly, we did not validate these methods in a clinical setting. This would have helped to determine sensitivity and specificity in colonized/infected patients and would have provided more clinical guidance in determining screening strategies. This was performed by two recent studies in Germany, 2017-2022 [21] and in the Netherlands, 2023 [11]. Both studies included patients that were screened because of a recent history of hospital admission abroad. *C. auris* was not detected by Heindel and colleagues who only used culture (CHROMagar, CC) for screening [21]. The study by Leonhard and colleagues did find seven cases of *C. auris* using the combined diagnostic approach of in-house qPCR followed by culture when qPCR was positive (CHROMagar, type unspecified) [11]. Only 1/7 PCR positive cases were also culture positive which could reflect an increased sensitivity of qPCR, or could indicate the presence of non-viable DNA and/or low level of colonization. This is clinically relevant since the degree of colonization might impact the risk of transmission as shown by Sansom and colleagues who observed a positive correlation between *C. auris* environmental contamination and the number of colonized body sites [20]. Third, we did not assess inter-operator repeatability because no triplicate testing was performed. This could have diminished the possibility of sampling error in samples with a low inoculum.

There is ongoing debate on how to implement screening and who should be screened in the Dutch low prevalence setting. From a laboratory-based perspective, Komorowski and colleagues [15] noted it is more cost-effective to screen only by culture but this study on cost-effectiveness did not take precaution equipment and isolation costs into account. The use of qPCR as a quick and reliable screening method prior to culture to detect *C. auris* carriage/infection limits isolation time and precautions. It depends on the local prevalence and setting (outbreak versus routine screening) whether this strategy is cost-effective. Cost-effectiveness studies in the clinical setting using a qPCR-based versus a culture-based approach are needed.

## CONCLUSION

Based on our results in mock-inoculated rectal specimens, the qPCR developed by Leonhard and colleagues [11]is the most reliable screening method to detect *C. auris* in lower inocula. Culture using CC and CCP plates is sensitive with inocula of ≥100 CFU/ml, but to increase specificity, the use of CCP agar and/or incubation at higher temperature is required. Future studies in the setting of routine screening should still compare qPCR combined with culture at 37°C and 42°C as the level of carriage remains elusive. Furthermore, the transmission risk should be assessed in patients that are only qPCR positive to establish whether a decreased sensitivity at 42°C for a low inoculum is clinically relevant.

## Data Availability

All data produced in the present study are available upon reasonable request to the authors

## REFERENCES

1. Liu, F., et al., Phylogenomic analysis of the Candida auris-Candida haemuli clade and related taxa in the Metschnikowiaceae, and proposal of thirteen new genera, fifty-five new combinations and nine new species. Persoonia, 2024. 52: p. 22–43.

2. World Health Organization, WHO fungal priority pathogens list to guide research, development and public health action. 2022: Geneva.

3. Welsh, R.M., et al., Survival, Persistence, and Isolation of the Emerging Multidrug-Resistant Pathogenic Yeast Candida auris on a Plastic Health Care Surface. J Clin Microbiol, 2017. 55(10): p. 2996–3005.

4. de Jong, A.W., et al., Performance of Two Novel Chromogenic Media for the Identification of Multidrug-Resistant Candida auris Compared with Other Commercially Available Formulations. J Clin Microbiol, 2021. 59(4).

5. Bentz, M.L., et al., Evaluation of CHROMagar Candida Plus for the detection of C. auris with a panel of 206 fungal isolates and 83 colonization screening skin-swabs. Microbiol Spectr, 2024: p. e0356423.

6. Marathe, A., et al., Utility of CHROMagar Candida Plus for presumptive identification of Candida auris from surveillance samples. Mycopathologia, 2022. 187(5-6): p. 527–534.

7. Du, H., et al., Candida auris: Epidemiology, biology, antifungal resistance, and virulence. PLoS Pathog, 2020. 16(10): p. e1008921.

8. Dennis, E.K., S. Chaturvedi, and V. Chaturvedi, So Many Diagnostic Tests, So Little Time: Review and Preview of Candida auris Testing in Clinical and Public Health Laboratories. Front Microbiol, 2021. 12: p. 757835.

9. OLM diagnostics. AurisID. Available from: https://olmdiagnostics.com/products/aurisid/.

10. Leach, L., Y. Zhu, and S. Chaturvedi, Development and Validation of a Real-Time PCR Assay for Rapid Detection of Candida auris from Surveillance Samples. J Clin Microbiol, 2018. 56(2).

11. Leonhard, S.E., et al., Proposal for a screening protocol for Candida auris colonization. J Hosp Infect, 2024.

12. Stavrou AA, G.v.d.E.B., Brouwer C, Boekhout T, Hagen F., Emerging Saccharomycotina yeast pathogens: Detection and susceptibility profiles. 2022.

13. Stevenson M. Package ‘epiR’ Tools for the Analysis of Epidemiological Data. 2024; Available from: https://cran.r-project.org/web/packages/epiR/epiR.pdf.

14. Borman, A.M., M. Fraser, and E.M. Johnson, CHROMagarTM Candida Plus: A novel chromogenic agar that permits the rapid identification of Candida auris. Med Mycol, 2021. 59(3): p. 253–258.

15. Komorowski, A.S., et al., Verification, Analytical Sensitivity, Cost-effectiveness, and Comparison of 4 Candida auris Screening Methods. Open Forum Infect Dis, 2024. 11(6): p. ofae017.

16. Dalben, Y.R., et al., Early Candida colonisation impact on patients and healthcare professionals in an intensive care unit. Mycoses, 2024. 67(8): p. e13786.

17. Li, Q., et al., Abundance interaction in Candida albicans and Candida glabrata mixed biofilms under diverse conditions. Med Mycol, 2021. 59(2): p. 158–167.

18. Kirkpatrick, W.R., et al., Growth competition between Candida dubliniensis and Candida albicans under broth and biofilm growing conditions. J Clin Microbiol, 2000. 38(2): p. 902–4.

19. Zhu, Y., et al., Laboratory Analysis of an Outbreak of Candida auris in New York from 2016 to 2018: Impact and Lessons Learned. J Clin Microbiol, 2020. 58(4).

20. Sansom, S.E., et al., Rapid Environmental Contamination With Candida auris and Multidrug-Resistant Bacterial Pathogens Near Colonized Patients. Clin Infect Dis, 2024. 78(5): p. 1276–1284.

21. Heindel, J., et al., Usefulness of screening for Candida auris colonisation in international patients admitted to a large university hospital. Mycoses, 2023. 66(2): p. 138–143.

