## Supplemental file for "Keep the hospital clean: diagnostic performance of ten different molecular and culture-based methods to detect *Candidozyma auris*"

**Table S1.** Strains used in this study

| Number | Name | Comment |
|---|---|---|
| CDC-AR0387 | *Candidozyma auris* | clade I, South-Asia |
| CDC-AR0388 | *C. auris* | clade I, South-Asia |
| CDC-AR0381 | *C. auris* | clade II, East-Asia; India |
| CDC-AR0382 | *C. auris* | clade II, East-Asia; India |
| CDC-AR0383 | *C. auris* | clade III, Africa |
| CDC-AR0384 | *C. auris* | clade III, Africa |
| CDC-AR0385 | *C. auris* | clade IV, South-America |
| CDC-AR0386 | *C. auris* | clade IV, South-America |
| CDC-AR1097 | *C. auris* | potential clade V, Iran |
| CBS 5149 | *Candidozyma haemuli* | Genetic mixture |
| CBS 10004 | *Candidozyma pseudohaemuli* | Genetic mixture |
| CBS 7798 | *Candidozyma duobushaemuli* | Genetic mixture |
| CBS 16213 | *Candidozyma khanbhai* | Genetic mixture |
| CBS 14366 | *Candidozyma vulturna* | Genetic mixture |
| CBS 562 | *Candida albicans* | Common mixture |
| CBS 604 | *Candida parapsilosis* | Common mixture |
| CBS 1920 | *Candida tropicalis* | Common mixture |
| CBS 138 | *Nakaseomyces glabratus (C. glabrata)* | Common mixture |
| CBS 13155 | *Pichia kudriavzevii (C. krusei)* | Common mixture |

**Table S2.** Molecular tests used in this study

| Name | | (Reporter)-Sequence-(Quencher) | Ref. |
|---|---|---|---|
| AurisID | Commercial: OLM Diagnostics, Braintree, United Kingdom. | Not available | [1] |
| *C. auris* AltoStar | Commercial: Altona Diagnostics, Hamburg, Germany | Not available | |
| *C. auris* screening assay | Commercial: Pathonostics, Maastricht, The Netherlands | Not available | |
| Leach PCR | In-house developed | Forward primer 5′-CAGACGTGAATCATCGAATCT-3′, Reverse primer 5′-TTTCGTGCAAGCTGTAATTT-3′, TaqMan-probe 5'-FAM-AATCTTCGCGGTGGCGTTGCATTCA-3IABkFQ-3′) | [2] |
| Leonhard PCR | In-house developed | Forward primer 5'-CCTGTTTGAGCGTGATGTCT, Reverse primer CGTGCAAGCTGTAATTTTGTG, TaqMan-probe 5'-FAM-CAACGCCACCGCGAAGATTG-3IABkFQ-3' | [3] |
| *C. haemuli* species complex | In-house developed | Forward primer 5'-TAAACGGCGGTCTTATCCTG-3', Reverse primer 5'-ATCTTCACCGCGAGTGCTAT-3', TaqMan-probe 5'-FAM-AGGATCCTAAGGTAGCGAAATTCATTGACA-3IABkFQ-3' | [4] |
| *C. auris* IDCARD | In-house developed | | [4] |

**Table S3.** Ct values for the samples from direct material versus after broth enrichment

| PCR | Direct: 50 CFU + common mix Mean | Broth: 50 CFU + common mix Mean | Direct: 50 CFU + genetic mix Mean | Broth: 50 CFU + genetic mix Mean |
|---|---|---|---|---|
| AurisID (OLM)* | 30.8 | 31.4 | 28.8 | 29.7 |
| AltoStar (Altona) | 36.5 | 37.8 | 35.5 | 36.4 |
| *C. auris* screening assay (PathoNostics) | 35.0 | 36.8 | 34.3 | 35.2 |
| Leonhard et al. qPCR | 34.2 | 35.9 | 34.2 | 34.2 |
| Leach et al. qPCR | 34.7 | 34.7 | 33.9 | 33.2 |
| *C. auris* IDCARD | 33.9 | 36.2 | 33.0 | 34.2 |
| *C. haemuli* complex qPCR | 33.8 | 36.4 | 29.9 | 34.5 |


1. OLM diagnostics. *AurisID*. Available from: https://olmdiagnostics.com/products/aurisid/.
2. Leach, L., Y. Zhu, and S. Chaturvedi, *Development and Validation of a Real-Time PCR Assay for Rapid Detection of Candida auris from Surveillance Samples.* J Clin Microbiol, 2018. **56**(2).
3. Leonhard, S.E., et al., *Proposal for a screening protocol for Candida auris colonization.* J Hosp Infect, 2024.
4. Stavrou AA, G.v.d.E.B., Brouwer C, Boekhout T, Hagen F., *Emerging Saccharomycotina yeast pathogens: Detection and susceptibility profiles.* 2022.
